# When can we stop wearing masks? Agent-based modeling to identify when vaccine coverage makes nonpharmaceutical interventions for reducing SARS-CoV-2 infections redundant in indoor gatherings

**DOI:** 10.1101/2021.04.19.21255737

**Authors:** Trevor S. Farthing, Cristina Lanzas

## Abstract

As vaccination efforts to combat the COVID-19 pandemic are ramping up worldwide, there are rising concerns that individuals will begin to eschew nonpharmaceutical interventions for preventing SARS-CoV-2 transmission and attempt to return to pre-pandemic normalcy before vaccine coverage levels effectively mitigate transmission risk. In the U.S.A., some governing bodies have already weakened or repealed guidelines for nonpharmaceutical intervention use, despite a recent spike in national COVID-19 cases and majority population of unvaccinated individuals. Recent modeling suggests that repealing nonpharmaceutical intervention guidelines too early into vaccine rollouts will lead to localized increases in COVID-19 cases, but the magnitude of nonpharmaceutical intervention effects on individual-level SARS-CoV-2 infection risk in fully- and partially-vaccinated populations is unclear. We use a previously-published agent-based model to simulate SARS-CoV-2 transmission in indoor gatherings of varying durations, population densities, and vaccination coverage levels. By simulating nonpharmaceutical interventions in some gatherings but not others, we were able to quantify the difference in SARS-CoV-2 infection risk when nonpharmaceutical interventions were used, relative to scenarios with no nonpharmaceutical interventions. We found that nonpharmaceutical interventions will often reduce secondary attack rates, especially during brief interactions, and therefore there is no definitive vaccination coverage level that makes nonpharmaceutical interventions completely redundant. However, the reduction effect on absolute SARS-CoV-2 infection risk conferred by nonpharmaceutical interventions is likely proportional to COVID-19 prevalence. Therefore, if COVID-19 prevalence decreases in the future, nonpharmaceutical interventions will likely still confer protective effects but potential benefits may be small enough to remain within “effectively negligible” risk thresholds.

## Introduction

Global vaccine rollout to combat the Coronavirus Disease 2019 (COVID-19) pandemic is well underway, with at least different seven vaccines approved for distribution by different countries (WHO 2021). In the U.S.A., where three vaccines have been approved for distribution (CDC 2021a), 24.8% of the population has been fully vaccinated against COVID-19 as of April 17^th^ 2021 (CDC 2021b). Despite ongoing vaccine rollouts, as of April 17^th^ 2021, there is an indication that COVID-19 cases are surging in some U.S. states (NY Times 2021). In spite of rising case numbers, several U.S. states have recently rescinded, or allowed to expire, policies mandating use of nonpharmaceutical intervention in public spaces, with seemingly no intention of reinstating them in the near future (State of Iowa 2021; State of Mississippi 2021; State of Texas 2021). Population-level epidemiological models of vaccine rollout effects on COVID-19 transmission suggest that discontinuing nonpharmaceutical intervention use early into the vaccination effort leads to a subsequent surge in COVID-19 cases and related hospitalizations and deaths (Gozzi et al. 2021; Moore et al. 2021).

The magnitude of nonpharmaceutical intervention effects on individual-level SARS-CoV-2 infection risk in fully- and partially-vaccinated populations is unclear. This information is crucial for identifying vaccination levels at which it would be appropriate to scale-back guidelines for nonpharmaceutical interventions, as it would allow governing bodies to base policies on concrete risk estimates. The United States Centers for Disease Control and Prevention (CDC) has updated guidelines on safe gathering protocols, recommending that groups of fully-vaccinated people can now safely interact amongst themselves, or with small groups of unvaccinated people at low risk for developing severe COVID-19, without utilizing any nonpharmaceutical Severe Acute Respiratory Syndrome Coronavirus 2 (SARS-CoV-2) transmission interventions (e.g., face coverings, 2-m social distancing, etc.) (CDC 2021c). However, the guidelines also recommend to continue avoiding medium to large gatherings, and the use of nonpharmaceutical interventions in public and when gathering with unvaccinated individuals. This caution stems from the incomplete knowledge of vaccine effectiveness across different populations, their effects on transmission, and the potential change on vaccine effectiveness caused by the emergence of new SARS-Cov-2 variants.

The problem with citing vaccination efforts as a justification for discontinuing nonpharmaceutical interventions is twofold. First and foremost, the majority of the U.S. population is not yet fully vaccinated (CDC 2021b), and therefore presumably has little-to-no immunity from SARS-CoV-2 infections. Secondly, while there is growing evidence that these vaccines reduce SARS-CoV-2 infection risk in addition to COVID-19 incidence, vaccines may not confer complete immunity or block transmission (Hall et al. 2021; Lipsitch & Kahn 2021; Yellen et al. 2021). Data suggest that the BNT162b2 mRNA vaccine (i.e., the vaccine developed by Pfizer-BioNtech) may be ≈ 72% effective at preventing laboratory-confirmed SARS-CoV-2 infections after a single dose, and ≈ 86-92% two weeks following the second dose (Hall et al. 2021; Yellen et al. 2021). Furthermore, this vaccine may reduce viral loads, a potential proxy for infectiousness, in infected individuals by 3-4 times (Levine-Tiefenbrun et al. 2021). Less information is available on the ability of the other two vaccines approved for U.S. distribution to reduce SARS-CoV-2 infections, but Lipsitch & Kahn (2021) do estimate that mRNA-1273 (i.e., the vaccine developed by Moderna and NIAID) can reduce individual-level infection risk by at least 61% following the first dose. Despite potentially-high infection-reduction efficacies, without vaccines that confer complete immunity from infection or prevent transmission from infectious individuals, it will be difficult to halt SARS-CoV-2 circulation in the population through vaccination efforts alone (Gozzi et al. 2021; Moore et al. 2021). Considering that most people also have yet to be fully vaccinated, guidelines that advocate phasing out nonpharmaceutical interventions during interpersonal interactions may be premature at this time.

In Farthing & Lanzas (2021), we described an agent-based model (ABM) for simulating indoor respiratory pathogen transmission. We previously used this model to quantify effects of nonpharmaceutical interventions on reducing SARS-CoV-2 transmission risk during an indoor superspreading event (Farthing & Lanzas 2021). Here, we use it to simulate SARS-CoV-2 transmission in indoor gatherings of varying durations, population densities, and proportional vaccination coverage. By simulating nonpharmaceutical interventions in some gatherings but not others, we were able to quantify the difference in SARS-CoV-2 infection risk when nonpharmaceutical interventions were used in conjunction with vaccination efforts, relative to scenarios with no nonpharmaceutical interventions. Using these data, we demonstrate how interested parties can easily estimate the potential reduction in SARS-CoV-2 infection risk attributable to nonpharmaceutical interventions, and try to answer the question: “at what point during vaccine rollout are gatherings without non-pharmaceutical measures safe?”

## Methods

We used the ABM we first described in Farthing & Lanzas (2021) to simulate the effect of increasing vaccination coverage and nonpharmaceutical interventions on SARS-CoV-2 transmission risk during indoor gatherings. The simulation input levels and parameter values we used are given in Table 1. We made the assumptions that any infectious individuals at gatherings would be asymptomatic because symptomatic people would consciously decide to stay away, and that no one with partial immunity exists within the group of attendees. Vaccinated people had a fixed probability of becoming completely immune to SARS-CoV-2 infection (Table 1), and those that did not become immune remained susceptible to infection (i.e, ‘all-or-nothing’ vaccine). Finally, we only simulated use of cloth face coverings, rather than notably more-effective masks like N95s, because we make the assumption that the majority of Americans have ready access to, and are more-likely to use cloth masks.

**Table 1.**
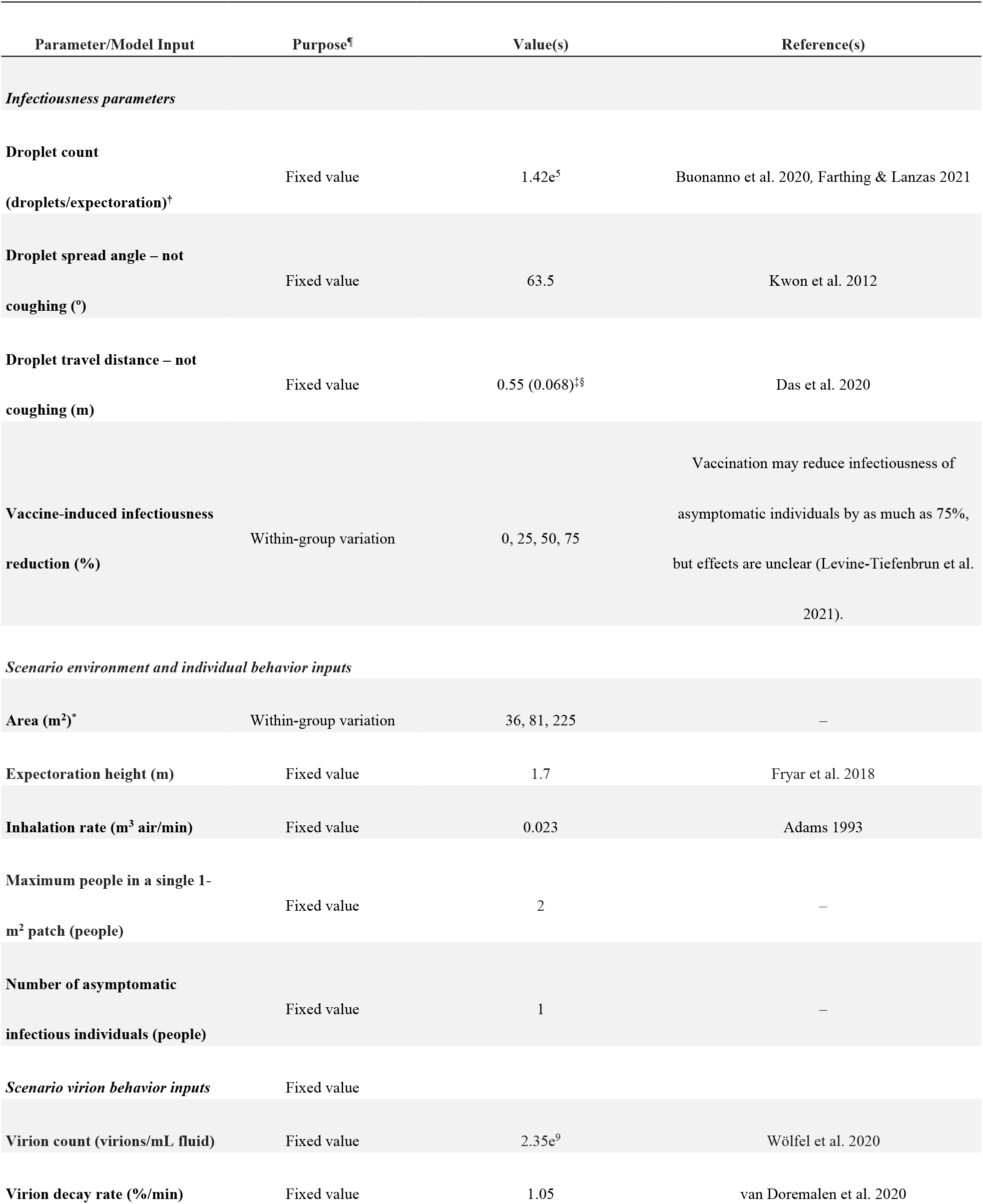
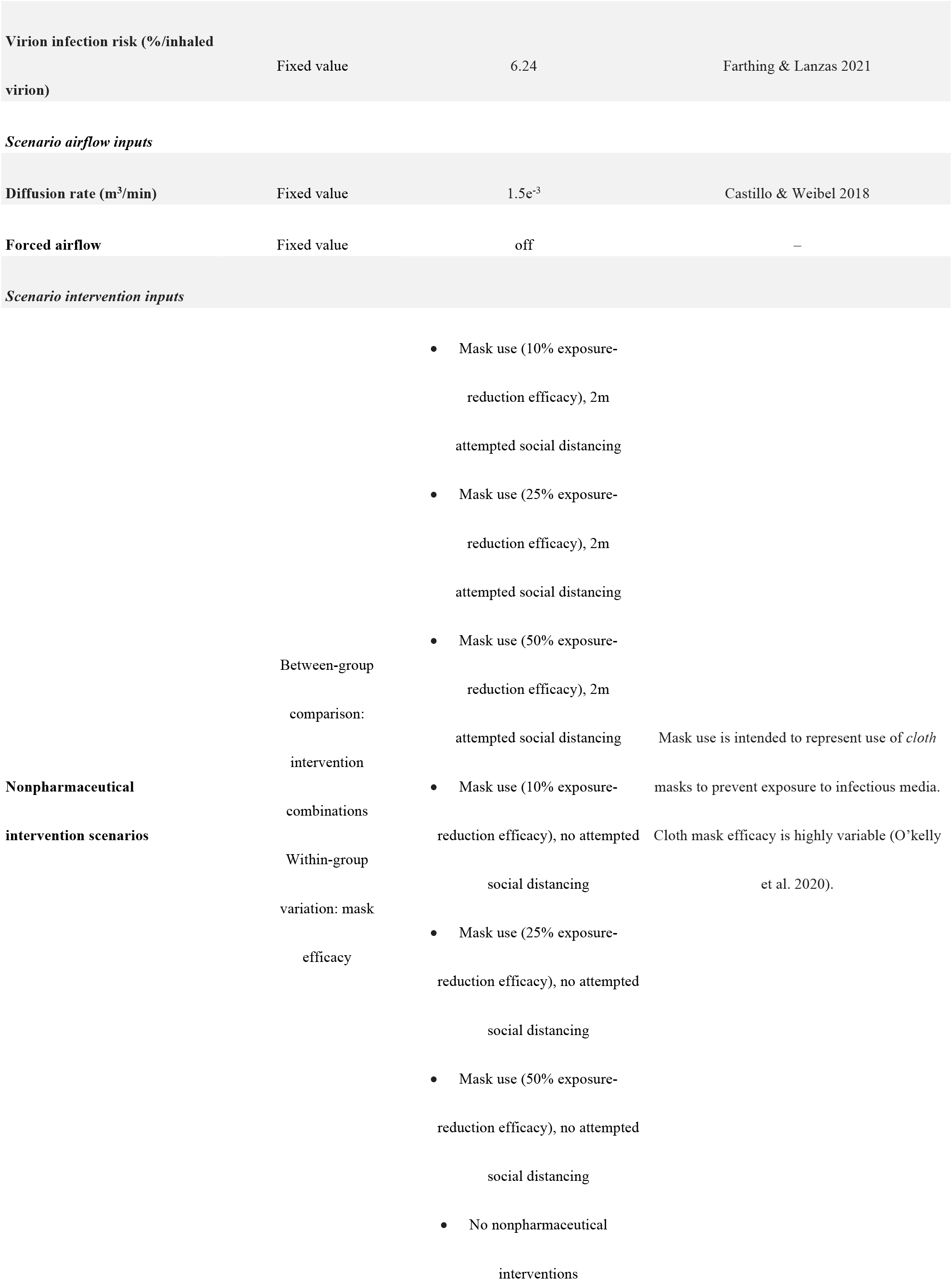
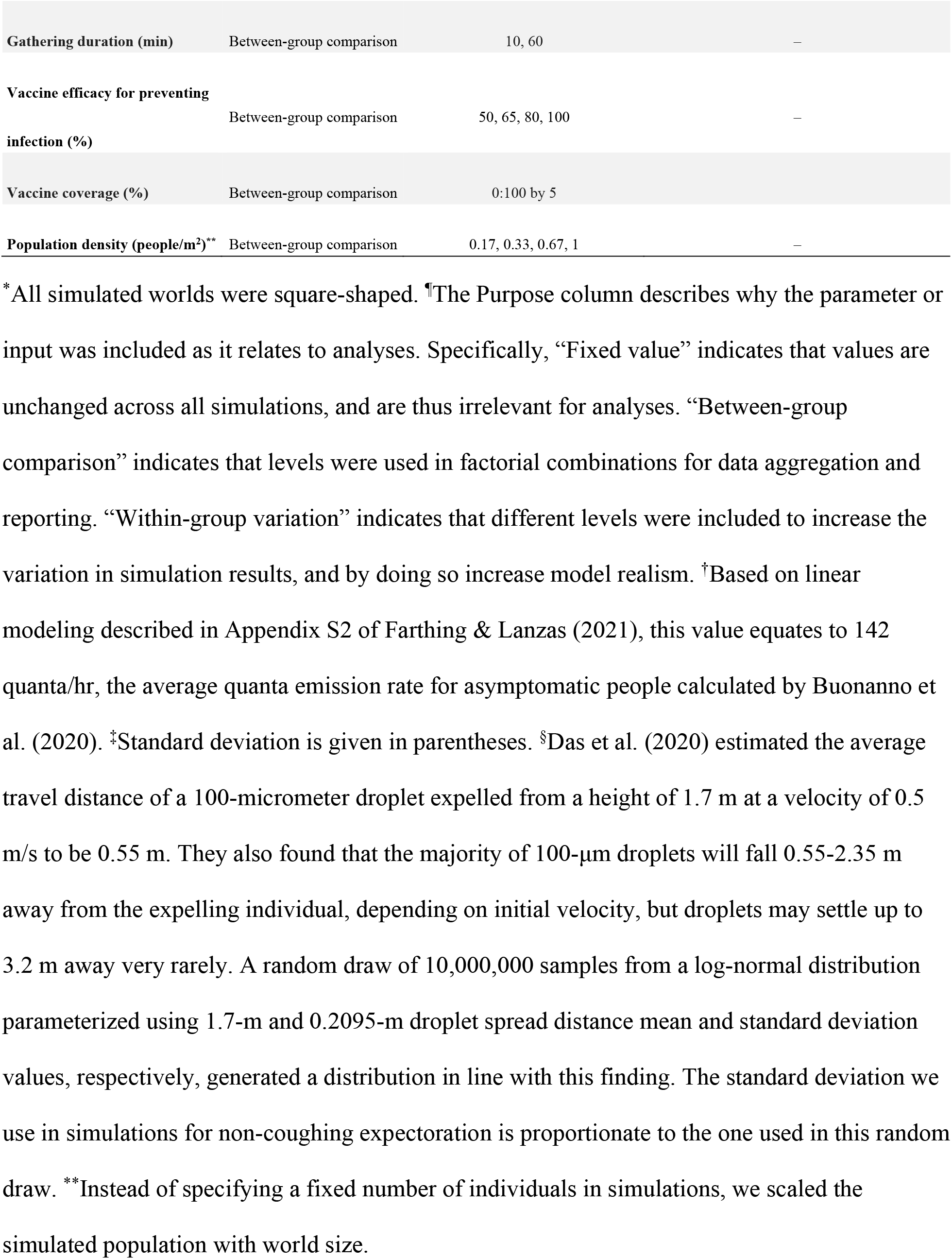
Model parameter and scenario-specific input descriptions for transmission simulations.

All simulations were carried out within the open-source modeling software, NetLogo (Ver. 6. 1. 1 – Wilensky 1999). We executed a factorial simulation run in the NetLogo BehaviorSpace using our specified input levels, and ran 200 simulations replicates of each parameter set combination when the nonpharmaceutical interventions were included and when they were not. We ran these factorial combination sets separately in order to save computation time as there were two inputs (i.e., mask efficacy, attempted social distance) that only changed when nonpharmaceutical interventions were simulated. We ultimately produced 1,612,800 simulations without nonpharmaceutical interventions, and 9,676,800 including them (i.e., 11,289,600 total simulations). We recorded the number of susceptible individuals infected in each simulation, and aggregated this information into a single data set prior to analysis.

We reported the mean probability of observing ≥ 1 successful infection event(s) and mean secondary attack rates in indoor gatherings when an asymptomatic person was also in attendance across factorial combinations of “between-group comparison” variables (Table 1). Secondary attack rates here were calculated by dividing the number of people that were infected at the gathering by the number of “healthy” people at the start of the gathering, and can also be considered to be the individual-level probability of a previously healthy attendee being infected at the gathering. To assess the difference between protection conferred by the simultaneous deployment of pharmaceutical and nonpharmaceutical interventions, versus use of only nonpharmaceutical interventions, we first smoothed the observed mean secondary attack rates (*μ*) by fitting them to a beta regression model with a fixed unknown precision parameter, *ϕ* using a logit link function to map (0,1) values (Ferrari & Cribari-Neto 2004). The specific model is given by:

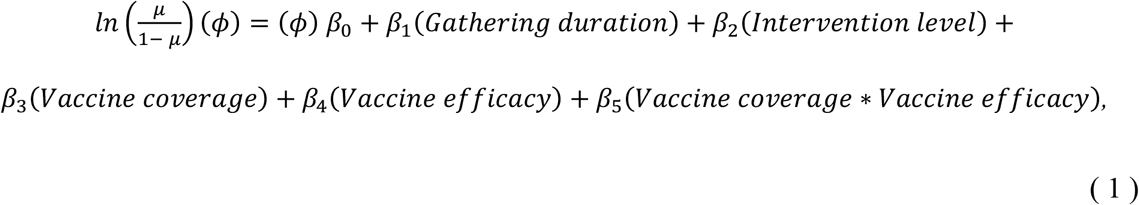

where “Intervention level” is a categorical variable containing the following mutually-exclusive levels: “cloth face masks & vaccination,” “cloth face masks & 2-m social distancing & vaccination,” and “vaccination only.” Additionally, “Vaccine efficacy” here refers to the ability of vaccines to induce complete immunity to infection. “Vaccine coverage” and “Vaccine efficacy” are given in terms of decimal percent, not percentage points (e.g., 0.1, not 10%).

Because beta regression models assume all dependent variable values fall between 0 and 1, we used the data transformation procedure described by (Cribari-Neto & Zeiles, 2010) to reconstruct our proportion data without these extremities prior to model fitting. We used the pseudo-R^2^ calculation procedure given by Ferrari & Cribari-Neto (2004) to assess the goodness of fit for our regression model.

After fitting our data, we used the regression model to predict the mean secondary attack rates during a 60-minute gathering with a single asymptomatic person in attendance across the complete factorial combination of covariate inputs described in Table 2. We report the difference between predicted values when all interventions (i.e., cloth face masks & 2-m social distancing & vaccination) are utilized, and predicted values assuming vaccinations are the only interventions. All analyses and plotting were carried out using functions from the “betareg” (Ferrari & Cribari-Neto 2004) and “ggplot2” (v. 3.3.2, Wickham 2016) R packages, respectively, in RStudio (v. 1.1.463, RStudio Team, Boston, MA) (RStudio Team 2018) running R (v. 3.6.2, R Foundation for Statistical Computing, Vienna, Austria) (R Core Team 2020).

**Table 2.** Covariate values used for prediction in our example.

| Covariate | Value(s) |
| --- | --- |
| Gathering duration | 60 min |
| Intervention level | cloth face masks & 2-m social distancing & vaccination<br>vaccination only |
| Vaccine coverage | 0:1 by 0.1 |
| Vaccine efficacy | 0.6, 0.8 |

## Results & Discussion

We found that the probability of ≥ 1 successful transmission event generally increased with population density (Fig. 1). This is unsurprising, as SARS-CoV-2 transmission in this ABM is highly sensitive to within-room population density (Farthing & Lanzas 2021). We observed that at low population densities and/or short-duration gatherings, the use of nonpharmaceutical interventions can significantly reduce the probability of successful transmission. Furthermore, it is clear that at low population densities, 2-m social distancing confers additional protective effects when used in conjunction with cloth face coverings, even during relatively-long duration gatherings. This is consistent with what we observed when we used the same ABM to directly compare the effectiveness of varied nonpharmaceutical interventions to prevent SARS-CoV-2 transmission during a superspreading event (Farthing & Lanzas 2021). We found that cloth face masks alone conferred few protective effects in long-duration gatherings.

**Figure 1.**
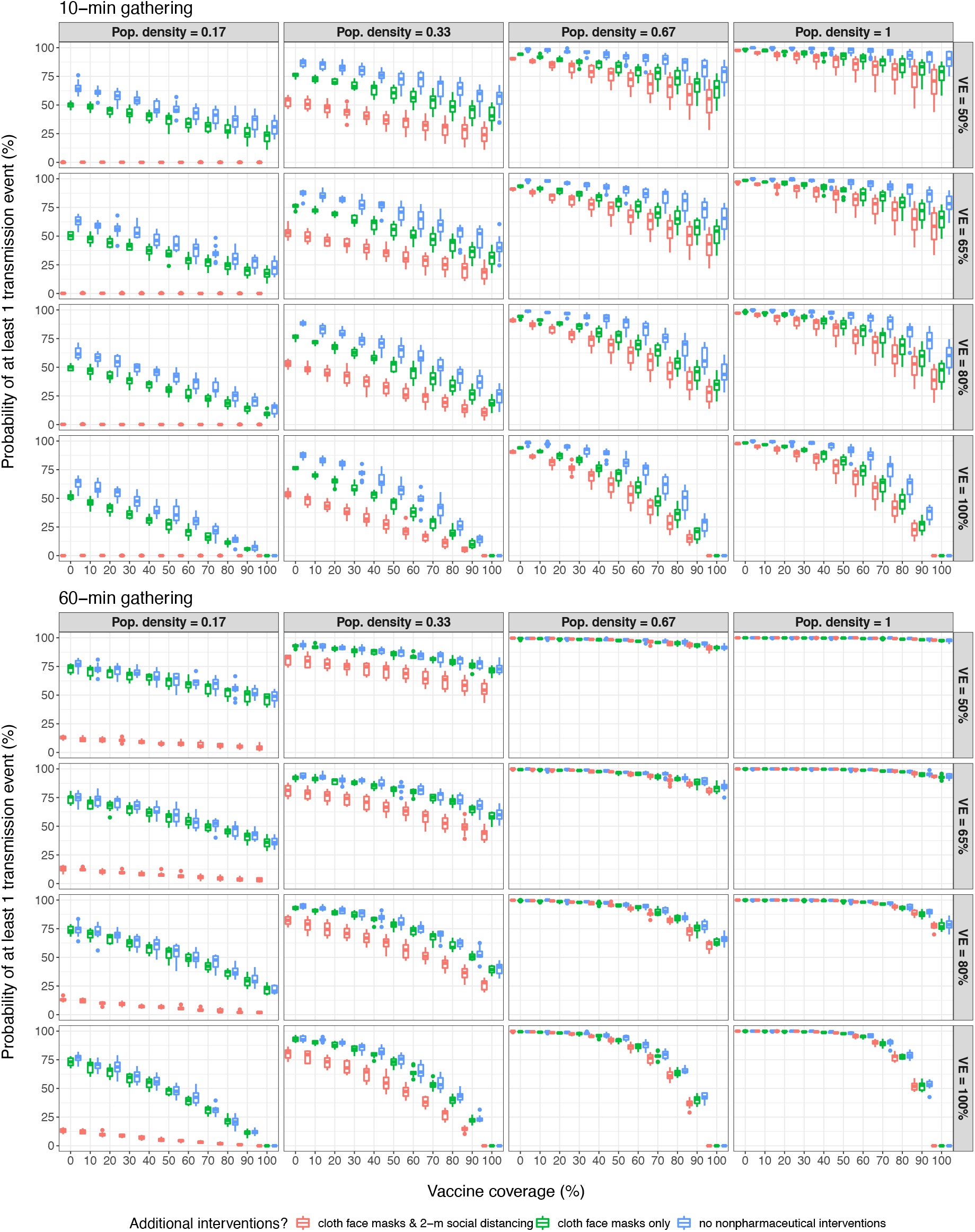
At low population densities and gathering duration limits, nonpharmaceutical interventions to prevent infection and elevated vaccination rates consistently decrease the probability of observing ≥ 1 successful SARS-CoV-2 transmission events in simulations.

The probability of transmission events occurring was unlikely to reach ≈ 0% outside of scenarios with low population density and multiple nonpharmaceutical interventions, or ≥ 95% vaccine coverage and vaccines that were 100% effective at preventing infections. Given that 1) current estimates place SARS-CoV-2 vaccine efficacies against infection between 60-90% (Hall et al. 2021; Lipsitch & Kahn 2021; Yellen et al. 2021), 2) historical precedence suggesting adult populations will fall well short of these high vaccination levels (Applewhite et al. 2020; CDC 2020), and 3) the difficulty government institutions have had enforcing nonpharmaceutical intervention policies (Jacobs & Ohinmaa 2020; Pedersen & Favero 2020), it is unlikely that these scenarios will be representative of average real-world gatherings. Moreover, in 60-min gathering scenarios, the probability of ≥ 1 successful transmission event occurring is relatively high even when gathering attendees utilize nonpharmaceutical interventions and most are vaccinated.

The probability that ≥1 SARS-CoV-2-positive individual is in attendance at a gathering can be calculated as

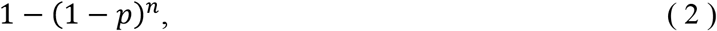

where *p* is the local COVID-19 prevalence, and *n* is the number of people at the gathering (Chande et al. 2020). The prevalence of infectious cases (*p*) can be highly uncertain because of the variable testing effort across time and space, but it can be estimated by assuming that any SARS-CoV-2-positive individuals are infectious at time of testing and will remain infectious for a given period of time. Additionally, ascertainment bias can be factored in. The probability that a given individual will be infected at a gathering is then

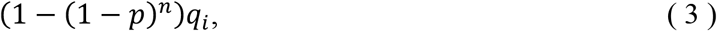

where *q*_i_ is the probability that individual *i* will be infected given exposure to an asymptomatic individual at the gathering. Effectively, what we report in Fig. 2 are estimates of *q*_”_ under different circumstances. Our findings suggest that cloth-based mask use, with or without 2-m social distancing, often does not confer significant protective effects during long-duration gatherings (Fig. 2), we have also shown that implementing these nonpharmaceutical interventions can reduce overall transmission probability (Fig. 1) and secondary attack rates (Fig. 2, Table 3) during brief interactions or gatherings with relatively-few people (e.g., fewer than 10 people, the limit for indoor and/or outdoor social gatherings enforced by some U.S. states (MultiState 2021)). This effectively means that strict guidelines for continued nonpharmaceutical intervention use will likely help to mitigate SARS-CoV-2 spread, and therefore COVID-19 incidence, for as long as these policies are in effect.

**Table 3.** Logit scale estimates associated with 1-unit increases in covariate values given by our beta-regression model. Wald 95% confidence intervals are given in parentheses. ^*^This is the reference level used to establish a baseline for binary dummy variables.

| Coefficient | Estimate | *p* |
| --- | --- | --- |
| Intercept | -3.786 (-3.857, -3.716) | – |
| $\phi$ | 28.899 (28.336, 29.462) | – |
| Gathering duration (min) | 0.012 (0.011, 0.012) | < 0.001 |
| Intervention level |  |  |
| *Cloth face masks & 2-m social distancing & vaccination\** | 0 (0, 0) | – |
| *Cloth face masks & vaccination* | 0.761 (0.737, 0.785) | < 0.001 |
| *Vaccination only* | 0.889 (0.866, 0.913) | < 0.001 |
| Vaccine coverage | 0.783 (0.660, 0.905) | < 0.001 |
| Vaccine efficacy | 0.385 (0.297, 0.472) | < 0.001 |
| Vaccine coverage X Vaccine efficacy | -2.652 (-2.816, -2.487) | < 0.001 |

**Figure 2.**
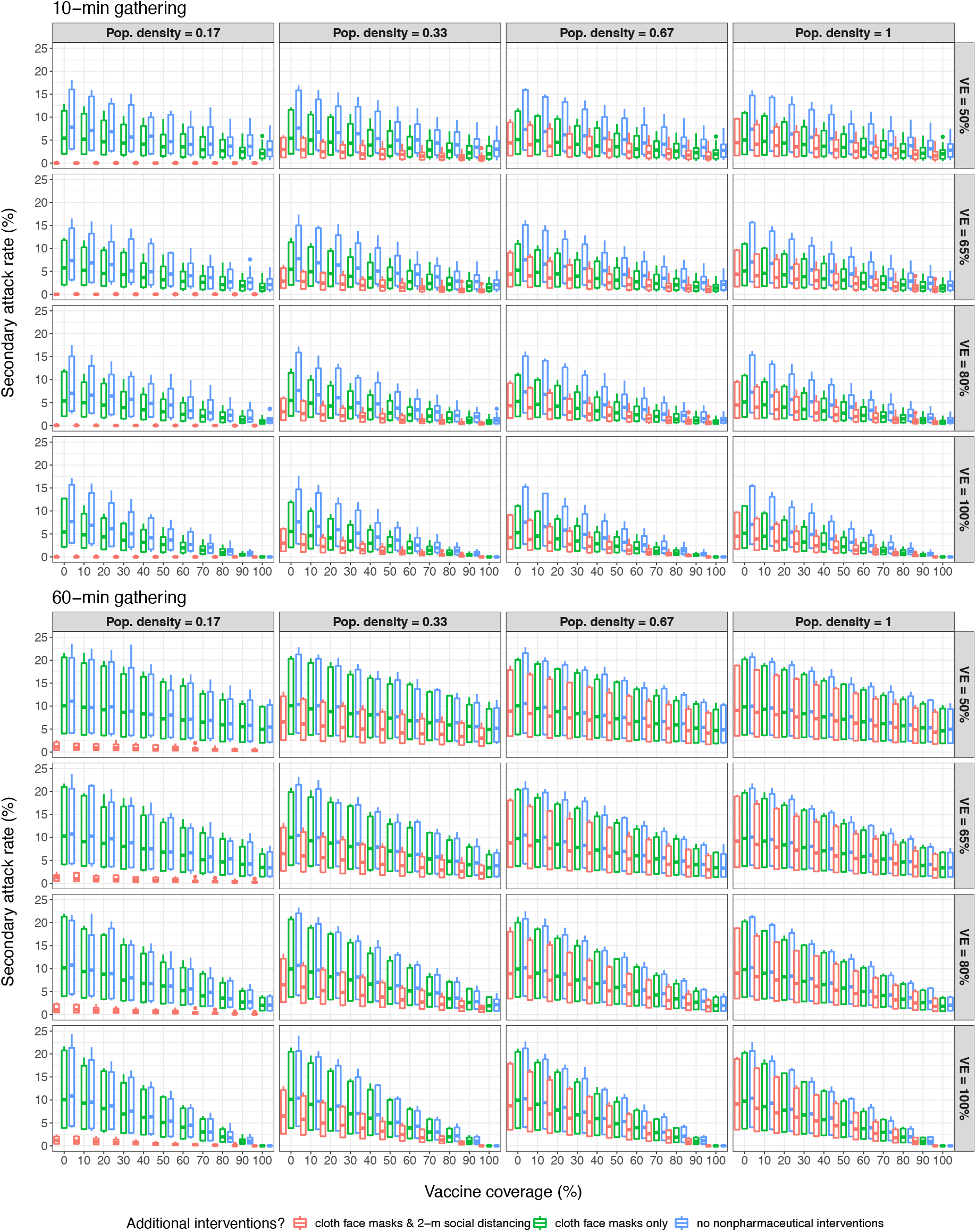
Mean secondary attack rates in simulations indicate substantial variability in risk.

As vaccine coverage increases, the question now becomes “how much elevated risk is acceptable in the absence of nonpharmaceutical interventions?” If we let 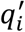 denote the probability that individual *i* will be infected given exposure to an asymptomatic individual at a gathering where no nonpharmaceutical interventions were in place, and 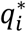 denote the probability that individual *i* will be infected given exposure to an asymptomatic individual at a gathering where some level of nonpharmaceutical interventions were in place, then the relative effect of nonpharmaceutical interventions on reducing infection risk is equal to

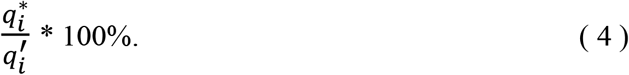

By quantifying covariate effects in our beta-regression model, we provide interested parties with a formula that can be used to quickly determine generalized 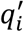 or 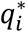 values, without the need for running a large number of simulations. Due to the logit link function we used, the mean secondary attack rates in our ABM simulations (μ) can be predicted using the equation

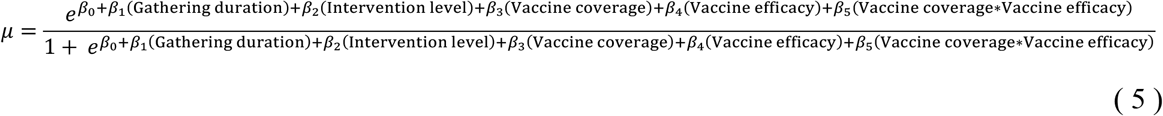

(Ferrari & Cribari-Neto 2004). Our regression model had a pseudo-R^2^ of 0.37. Given the number of stochastic processes in our ABM and the variability purposely introduced into simulations (Table 1), we believe the explanatory power of the model is acceptable for our purposes here.

Assuming mean population-level vaccine efficacies of 60% and 80%, which we believe are conservative estimates for U.S.-approved vaccine efficacies, our regression model consistently predicts that secondary attack rates decrease by 55-58% when attendees utilize cloth masks and 2-m social distancing, regardless of gathering duration (Fig. 3). However, it is important to reiterate that here we estimate the probability or infection given contact with an infectious individual at a gathering (*q*_i_) and comment on the relative risk difference attributable to intervention use. This should not be confused with the absolute risk of becoming infected at a gathering (*see* Equation 3). We demonstrate the difference in Figure 4, which is a simplistic example intended to show that even at relatively high COVID-19 prevalence levels, 20 people gathering indoors for 60 minutes have a substantially-lower individual-level risk of SARS-CoV-2 infections than is suggested by *q*_i_ alone. Though predicting intervention effects on community-level COVID-19 prevalence and infection-related events (e.g., symptom-onset, mortality, or hospitalization) is outside the scope of our model, our simulations do suggest that secondary attack rates are negatively correlated with vaccine coverage. Given that we expect local COVID-19 prevalence to eventually follow similar trends (Gozzi et al. 2021), the relative impact of nonpharmaceutical interventions on infection risk reduction will likely decrease over time as vaccine rollouts continue.

**Figure 3.**
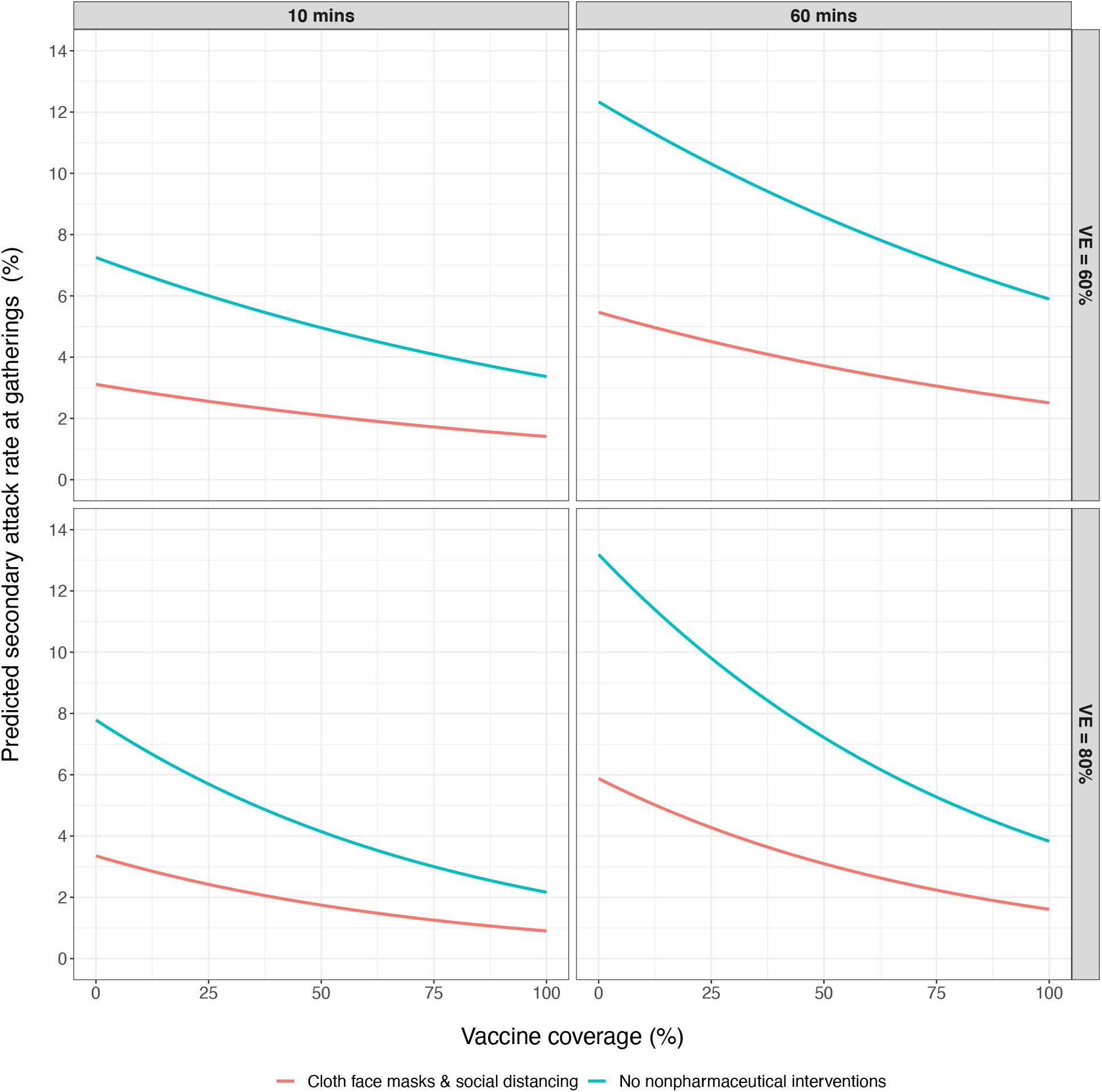
Predicted secondary attack rates suggest that the combination of cloth face masks and 2-m social distancing during indoor gatherings of varying durations consistently reduces secondary attack rates by 55-58%. This effect was only modeled for vaccine efficacies of 60% and 80%.

**Figure 4.**
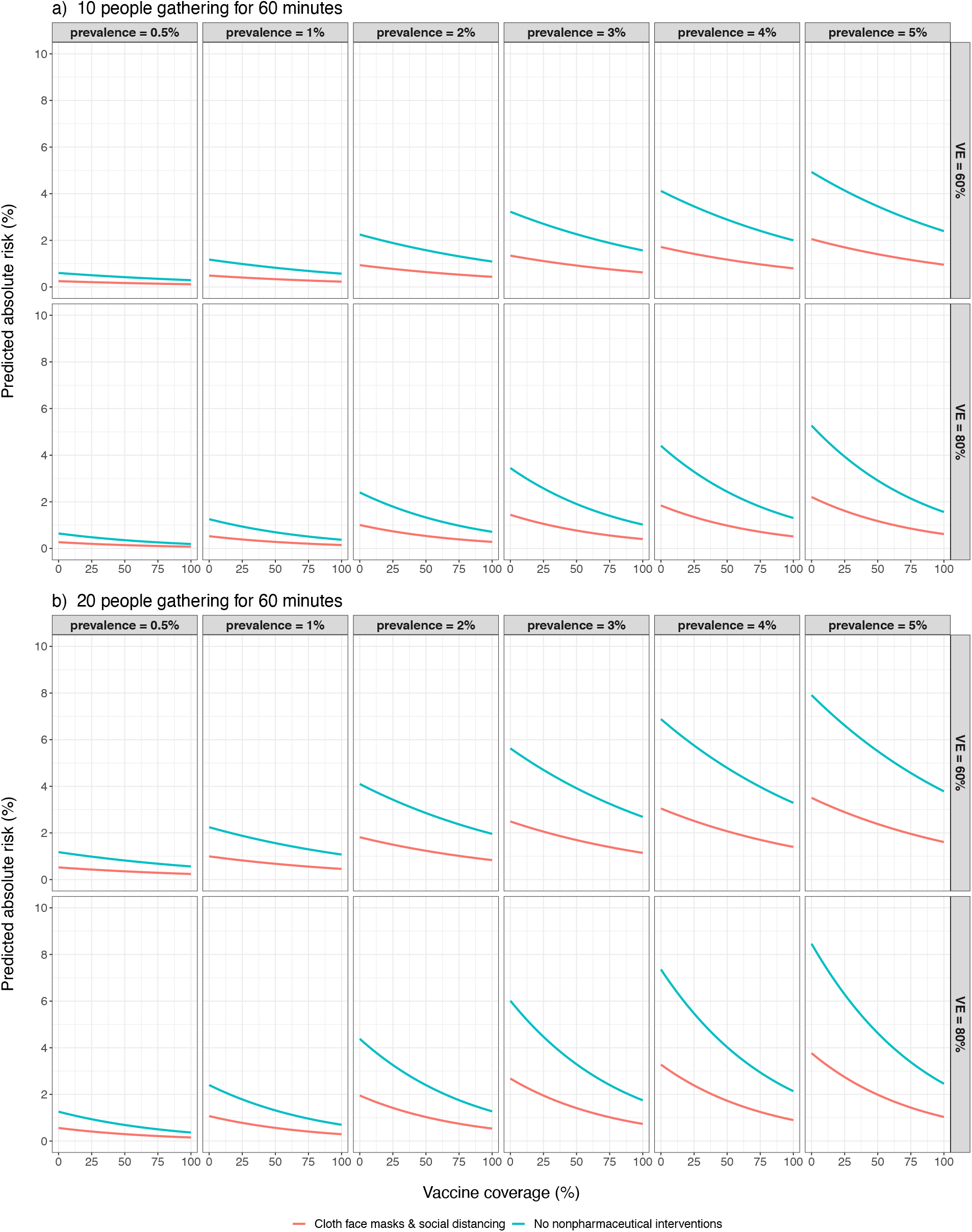
Estimated absolute risk of being infected with SARS-CoV-2 during 60-minute gatherings of varied sizes. Estimates were obtained by plugging Figure 3 predictions into Equation 3 with fixed COVID-19 prevalence and n values. a) Absolute risk of SARS-CoV-2 transmission given that 10 people attend the gathering. b) Absolute risk of SARS-CoV-2 transmission given that 20 people attend the gathering.

In addition to being unable to comment on community-level infection metrics, there are a few other limitations associated with our results that we must acknowledge. Aside from the ABM design limitations outlined in Farthing et al. (2021), we make a number of assumptions in our simulations. Most of these assumptions are directly tied to our parameter space detailed in Table 1, and include such things as: in simulated gatherings only one asymptomatic individual was in attendance, no individuals wear masks with exposure-reduction efficacies > 50% and therefore we are not simulating the use of N95 or similar masks, and there is no simulated forced-air ventilation or infectious individuals that produce superspreader-level of contaminated aerosols (e.g., 970 quanta (Miller et al. 2020)). Additionally, we do not simulate activity-specific behaviors and individuals in our simulations were unmoving. Finally, we based the infectiousness of asymptomatic individuals on the estimate given by Buonanno et al. (2020) (i.e., 142 quanta/hr), and to relate this estimate to ABM parameters we used the linear model described in Farthing et al. (2021). However, this parameterization procedure may have over-inflated virion transmissibility in certain scenarios because quanta-estimates are room-size specific, and the Farthing et al. (2021) linear model was based on simulations of gatherings within a relatively large room. In short, our results must be viewed through the lens of simulated world parameters and behaviors, and likely will not wholly reflect all variability that may exist in real-world transmission events. This is very common for ABM-based studies however, and we feel that our model is sufficiently accurate to highlight general trends in indoor SARS-CoV-2 transmission and infection risk.

## Conclusions

We found that nonpharmaceutical interventions will often reduce secondary attack rates, especially during brief interactions, and therefore there is no definitive vaccination coverage level that makes nonpharmaceutical interventions completely redundant. However, the beneficial effect on absolute SARS-CoV-2 infection risk reduction conferred by nonpharmaceutical interventions used during indoor gatherings is likely proportional to COVID-19 prevalence.

Therefore, if U.S. COVID-19 prevalence decreases in the future, nonpharmaceutical interventions will likely still confer protective effects, but any potential benefits may be small enough to remain within “effectively negligible” risk thresholds.

## Data Availability

We first made our ABM publicly available for download in Farthing et al. (2021). The current iteration can be downloaded from the Lanzas lab github repository at https://github.com/lanzaslab/droplet-ABM.

## Acknowledgments

This work was partially supported by CDC U01CK000587-01M001 and R35GM134934.

## Author Contributions

Trevor Farthing led the model creation, data analysis, and manuscript writing, but both authors conceived the ideas presented herein, contributed to model development and writing efforts, and gave final approval for publication. Cristina Lanzas secured the funding.

## Data availability

We first made our ABM publicly available for download in Farthing et al. (2021). The current iteration can be downloaded from the Lanzas lab’s github repository at https://github.com/lanzaslab/droplet-ABM.

